# Informal support networks and their role in community safety and mental well-being among racially and ethnically minoritised groups in London: a qualitative study

**DOI:** 10.64898/2026.02.17.26346389

**Authors:** Lewis Benjamin, Dorothy Williams, Zara Asif, Sanchika Campbell, Daniella Mousicos, Rebecca Rhead, Nathan Stanley, Hanna Kienzler, Stephani Hatch

**Author notes:** **Corresponding author: Lewis Benjamin**. **Ethics statement** This study was approved by the King’s College London Research Ethics Committee for Psychiatry, Nursing and Midwifery; approval number: HR/DP– 21/22-26357.

## Abstract

**Background:** Community safety is a key determinant of mental well-being, yet racially and ethnically minoritised communities in the UK often face higher exposure to violence alongside barriers to formal protection and support. In these contexts, informal support networks may play a critical role in shaping how safety is experienced and how distress is managed. Although such networks are widely recognised as protective for mental well-being, there is limited qualitative research examining how they operate in relation to community safety in settings shaped by structural inequality. This study explores how informal support networks influence experiences of community safety and mental well-being among racially and ethnically minoritised groups in South East London.

**Methods:** This qualitative study draws on semi-structured interviews (n = 31) with racially and ethnically minoritised participants aged 16+ living or working in Lambeth and Southwark [South East London]. Using a co-produced qualitative design, community consultations informed the development of interview topics. Interviews explored informal support networks, experiences of community safety and their intersections with mental well-being. Audio-recorded interviews were transcribed verbatim and analysed using inductive thematic analysis.

**Results:** Four themes were identified: (1) experiences of community safety and their mental health impacts; (2) gendered experiences of safety and responsibility; (3) formal support and its barriers; and (4) community and peer-led initiatives as a response to institutional distrust.

**Conclusion:** Informal support networks are central to everyday safety and emotional well-being, yet they cannot substitute for adequately resourced, culturally informed public provisions. Strengthening public infrastructure must involve meaningful collaboration with trusted community networks and address the intersectional needs of racially and ethnically minoritised groups.

## Introduction

Community safety, understood as the conditions that enable people to feel and be safe in their neighbourhoods, is widely recognised as a determinant of mental well-being (Bennetts et al., 2017). Although recorded crime represents one aspect of safety, broader contextual factors, such as social cohesion and neighbourhood stability, play a critical role in shaping everyday experiences of safety (Bennetts et al., 2017). Persistent exposure to unsafe environments is consistently associated with elevated levels of psychological distress (Laurencin & Walker, 2020). Ensuring community safety is therefore fundamental to protecting mental well-being at both individual and collective levels.

However, experiences of safety are not evenly distributed across populations. In the United Kingdom (UK), racially and ethnically minoritised groups — positioned through structural processes of racialisation and discrimination — face disproportionate exposure to violence and insecurity (Burnett, 2013). Exposure to violence within racially and ethnically minoritised communities is multifaceted, arising from both direct interpersonal harms and broader structural inequalities that sustain insecurity.

Interpersonal violence, including assault and abuse, carries immediate and enduring psychological consequences and is associated with elevated levels of trauma and distress (Romito et al., 2005). These harms are compounded by structural conditions such as neighbourhood disinvestment and limited access to early mental health support, which intensify chronic stress and reinforce feelings of unsafety (Laurencin & Walker, 2020). Together, these interpersonal and structural exposures create cumulative risks for poor mental well-being among racially and ethnically minoritised groups.

These inequalities are evident in the UK context. Racially motivated hate crimes continue to constitute the majority of recorded hate crimes in England and Wales (ONS, 2024). Racially and ethnically minoritised communities are also disproportionately concentrated in under-resourced neighbourhoods characterised by higher crime levels and reduced access to mental health services (The Health Foundation, 2024). Evidence from the South East London Community Health (SELCoH) study illustrates this mental health burden, demonstrating higher exposure to community violence and increased prevalence of post-traumatic stress disorder and depression among racially and ethnically minoritised populations (Kadra et al., 2014).

Crucially, these patterns of exposure intersect with barriers to formal care systems. Racially and ethnically minoritised communities frequently encounter obstacles when accessing mental health and safeguarding services. Anticipated and experienced racism within healthcare settings, alongside mistrust shaped by discriminatory policing and institutional neglect, can deter help-seeking and compound psychological distress (Williams & Mohammed, 2009; Hatch et al., 2016). The conditions of community safety and mental well-being are thus determined by the extent of harm people face and by how accessible protective care institutions are. Where formal care systems are perceived as inaccessible or untrustworthy, alternative sources of support may assume greater importance in sustaining everyday safety and managing distress.

In contexts of limited institutional trust, informal support networks therefore play a central role in responses to insecurity and emotional distress. Family members, friends, neighbours, faith groups, and community-led initiatives frequently serve as first points of contact when safety is threatened or mental well-being is strained (Memon et al., 2016). Through these networks, individuals may access emotional reassurance and culturally grounded forms of understanding that are not always available within statutory services. Research indicates strong social ties are associated with more favourable mental health outcomes (Smyth et al., 2015). In urban contexts where institutional trust is low, such informal networks may also support collective vigilance and shared forms of protection in everyday life, reinforcing both perceived safety and overall mental well-being.

Yet these networks are not without constraints. Strong internal ties may limit disclosure of sensitive experiences, particularly gender-based violence and mental health, or reinforce norms that discourage engagement with external services (Misra et al., 2021). Moreover, informal networks may become overburdened in contexts of chronic insecurity, acting as substitutes for formal systems that should be providing safety and mental health support (Villalonga-Olives & Kawachi, 2017). Their role must therefore be understood as relational and context-dependent, in that they may be both protective and potentially constrained by the structural conditions in which they operate.

Experiences of safety are further shaped by intersecting social identities, including gender, race, migration status, and socio-economic position. Violence against women and girls (VAWG) illustrates how overlapping social inequalities produce vulnerability (Cho et al., 2013). Racially and ethnically minoritised women face heightened exposure to gender-based violence, including domestic abuse, sexual harassment, and honour-based violence (Clark et al., 2008). The psychological consequences of VAWG, including long-term trauma and increased distress (Petrosky et al., 2017), are often compounded for racially and ethnically minoritised women by the limited availability and accessibility of culturally responsive services, despite evidence that culturally adapted trauma care can significantly improve outcomes (Benjamin et al., 2025).

Moreover, gender shapes the internal organisation of community care. Emotional labour and the coordination of informal support frequently fall disproportionately on women, particularly where institutional support is weak (Evans & Feder, 2016). Informal networks thus sustain everyday safety, but often through gendered distributions of responsibility that carry their own psychological costs.

### Theoretical framework

This study is informed by the concepts of structural violence, structural racism, and intersectionality, which together provide a robust framework for understanding how inequalities in safety and mental well-being are produced and sustained.

*Structural violence* refers to the ways in which social and economic structures systematically restrict access to essential resources and care, producing avoidable harm (Guillot-Wright et al., 2024). In the context of community safety and mental health, structural violence operates through material conditions such as neighbourhood disinvestment and limited service provisions, as well as through institutional practices that offer uneven support. These conditions embed inequality into everyday life and contribute to psychological distress (Murray et al., 2026).

*Structural racism* conceptualises racism as a system of power embedded in institutions and policies that produces unequal exposure to risk and differential recognition of needs across racially and ethnically minoritised groups (Bailey et al., 2017). It shapes who is exposed to violence, whose safety concerns are taken seriously, and who can obtain adequate mental health support. In domains such as housing, policing, and health and social care, racialised hierarchies create patterned disparities in exposure to harm and access to care (Murray et al., 2026).

*Intersectionality* provides a framework for analysing how multiple systems of oppression interact to produce distinct experiences of disadvantage (Cho et al., 2013). Social categories, including gender, race, class, migration status, and disability are mutually constitutive, creating compounded vulnerabilities that cannot be understood in isolation. This perspective is critical for understanding experiences of violence and safety, as well as the unequal access to support, across racially and ethnically minoritised populations and social contexts.

Together, these frameworks position informal support networks as relational infrastructures embedded within broader systems of racialised inequality. They guide the analysis by attending both to the protective roles of informal support and to the systemic constraints that shape their availability and effectiveness.

### The present study

Although informal networks are increasingly recognised as important determinants of mental health, limited research has examined how informal support networks operate within racially and ethnically minoritised communities facing sustained insecurity and low institutional trust. In particular, there is a lack of empirical work exploring how safety is negotiated in everyday life, how informal networks relate to mental well-being, and how these processes are shaped by gendered dynamics. This qualitative study therefore examines how informal support networks influence experiences of community safety and mental well-being among racially and ethnically minoritised groups in South East London. Specifically, it explores:

1. How do informal support networks influence perceptions of safety and mental well-being among racially and ethnically minoritised groups in South East London?
2. In what ways do these networks address the gendered experiences of safety, particularly violence against women and girls?
3. How do they complement or substitute formal mental health support services?

By situating informal support within structures of structural violence, structural racism, and intersecting inequalities, this study advances mental health research by demonstrating how informal systems of care shape lived experiences of safety and mental well-being alongside formal services. By doing so, the study identifies implications for strengthening culturally informed responses to community safety and mental well-being.

## Methods

### Study design and context

This paper is based on a sub-study within a larger research programme titled: The CONtributions of social NEtworks to Community Thriving (CONNECT), funded by the Economic and Social Research Council (Reference: ES/S012567/1). The CONNECT study was a mixed-methods research programme exploring how social networks and social capital promote or hinder community thriving and mental well-being among racially and ethnically minoritised groups in South East London, specifically in the boroughs of Southwark and Lambeth.

Of all 333 local authorities in England, Lambeth and Southwark are the 20^th^ (Lambeth) and 15^th^ (Southwark) most ethnically diverse (Impact on Urban Health, 2023). A quarter of Lambeth’s population live in poverty, and over one third of Southwark residents live in areas with the highest levels of deprivation in England (Impact on Urban Health, 2023b), depicting vast inequalities that impact health and well-being. Lambeth is consistently ranked among the ten most high-crime boroughs within Greater London. In 2025, the borough recorded an overall crime rate of 99 incidents per 1,000 residents. This figure is significantly higher, approximately 27%, than the London-wide average of 78 crimes per 1,000 residents (CrimeRate, 2025a). Similarly, the neighbouring borough of Southwark also falls within the top ten boroughs in London for overall crime. In 2025, Southwark reported a crime rate of 88 incidents per 1,000 residents, approximately 13% above the London average (CrimeRate, 2025b), highlighting the social vulnerability additionally faced by residents across both boroughs.

The CONNECT study examines how social capital and social networks influence mental health and well-being across four interconnected priority areas: community safety; violence against women and girls; skills and employment; and food security. These topics were identified as priority concerns through initial community consultations and later refined via participatory discussions with local councils and community members. The study seeks to understand how social connections, both informal and institutional, serve as protective resources that can support individual and collective resilience within racially and ethnically minoritised communities. Methodologically, the overarching study was grounded in a Participatory Action Research framework (Baum et al., 2006), and combined qualitative interviews, quantitative analyses of healthcare datasets, and creative methods such as Photovoice and community mapping workshops. Further details of methodology are outlined in a previously published CONNECT research protocol (Gnan et al., 2023).

This paper focuses on the qualitative strand of the CONNECT study which involved semi-structured interviews, where participants selected one of two topics, including (1) food insecurity, employment, and mental well-being and (2) community safety, including VAWG, and mental well-being. This paper specifically focuses on data derived from the community safety and mental well-being interviews.

This sub-study used a co-produced qualitative design, grounded in collaborative inquiry and shared decision making (Graham et al., 2022). Partner organisations, such as those from local government, health providers, and third sector organisations, were involved from the outset and contributed to the conceptualisation and design of the study. Their ongoing involvement helped pivot aspects of the research to reflect changing needs, such as identifying trusted local hubs for participant recruitment and supporting the development of the interview topic guide. Their insights drew on years of grassroots engagements with the local community, including pre-existing consultations conducted by Lambeth and Southwark Councils (local government for the London boroughs) around community safety and mental well-being.

### Peer researchers

We recruited eight peer researchers to the study team. Selection criteria included being over the age of sixteen, belonging to a racially minoritised group, residing in the study area, an interest in qualitative methods and community research, and availability for training and fieldwork. Peer researchers played a significant role in shaping the study, and their expertise was instrumental in ensuring the study was relevant to the needs of participants. Due to the sensitive nature of exploring community safety and mental well-being, peer researchers underwent a structured training programme to equip them with qualitative research skills. Training was delivered through an online toolkit developed by members of the research team (https://www.futurelearn.com/courses/research-methods-a-practical-guide-to-peer-and-community-research) and supplemented by access to formal research training opportunities and ongoing supervision. Peer researchers co-produced protocols for recruitment, including developing creative recruitment videos, delivering training, conducting interviews, as well as contributing to reflexivity in research, such as keeping reflexive practice logs throughout the research process. Like partner organisations, their involvement supported the cultural grounding of the study and strengthened its methodological and ethical integrity. Peer researchers were compensated financially for their time.

### Positionality statement

We remain critically aware that this research was conducted within a university context, which shapes how we understand and engage with issues of community safety and the lived experiences of racially minoritised groups. We acknowledge that our positionings bring with them forms of power and privilege, especially in relation to communities that have been underserved without active involvement in research frameworks. We approached this project with a commitment to reflexivity. We engaged in regular critical reflection on the ethical and social dimensions of the research, and we view this reflexivity as integral to co-producing work that is methodologically sound. We collaborated with peer researchers and community stakeholders, working to ensure their insights informed the research design, data collection, analysis, interpretation, and the dissemination of findings. This collaborative approach aimed to build trust and challenge extractive research practices (Orr & Bennett, 2009) and ultimately support more relevant understandings of community safety and mental well-being.

### Recruitment

Participants were recruited from racially minoritised groups living or working in South East London. Eligibility criteria included being aged 16+, self-identifying as from a racialised minority background, and living or working in Southwark or Lambeth within the past five years, to ensure participants could speak to recent experiences relevant to the study aims. Participants were recruited through a mix of snowball and purposive sampling methods. Study advertisements were distributed through our partner organisations’ networks through flyers and social media. Members of the research team also compiled a database of relevant community and grassroots organisations that aligned with the study aims and priority areas. Recruitment through these organisations took place both in person (e.g., dissemination of flyers or engagement with people in public spaces including libraries and community organisations), as well as online. Recruitment took place between June 2023 and June 2024. Ongoing discussions on data saturation occurred between peer researchers and the wider research team, with an agreement that thematic saturation was reached after conducting thirty-one interviews.

### Participants

The majority of participants were female (n=24); five were male, one did not disclose their gender, and one response was missing. The largest age groups were 35–44 and 45–54 (n=8 each), followed by 16–24 (n=7), 25–34 (n=4), and 65+ (n=3); one age response was missing. Most participants identified as Black (n=22, including Black African and Black Caribbean). Others identified as Chinese (n=3), and one each as Bangladeshi, Indian, Mixed heritage, Pakistani, or White Other; one ethnicity was undisclosed, and one ethnicity response missing. Most identified as heterosexual (n=19), five as bisexual, one as gay or lesbian, two as “other,” three did not disclose, and one response was missing. The sample included 13 employed and 17 unemployed individuals, with one missing employment response.

### Data collection

Semi-structured interviews were conducted by both peer researchers and members of the academic research team. Interviews were conducted either online or in person in line with participants’ preferences. Participants provided informed consent before participation in the study and were made aware they were able to withdraw from the study at any time. Given the potentially sensitive nature of the topics, participants were signposted to appropriate local support services following participation. Interviews lasted up to sixty minutes and were audio-recorded and transcribed verbatim, with identifying names removed. Participants were compensated financially with a shopping voucher for taking part.

### Data analysis

A thematic analysis approach (Clarke & Braun, 2017) was used to analyse interview data. Transcripts were anonymised and then read multiple times for familiarisation. Two researchers (LB and DM) then systematically coded data using an inductive approach, identifying segments of text related to informal support networks, community safety, and their intersections with mental well-being. Following initial coding, both researchers met to compare and refine codes. Broader thematic categories were developed iteratively and reviewed alongside the full dataset. Emerging themes were then discussed with the wider CONNECT team to validate relevance and refine interpretations. Finally, themes were developed into a coherent narrative with references to participant quotations to depict important findings.

## Results

Four themes emerged from the data: (1) experiences of community safety and their mental health impacts, (2) gendered experiences of safety and responsibility, (3) formal support and its barriers, and (4) community and peer-led initiatives as a response to institutional distrust. Alongside participants’ extracts, pseudonym names and age brackets are reported to protect confidentiality.

### Experiences of community safety and their mental health impacts

Participants’ accounts demonstrated that safety was understood as a multidimensional experience, shaped by everyday interactions and neighbourhood cohesion. Safety was often described as a felt sense of reassurance and belonging within one’s immediate environment. Conversely, experiences of violence, or the anticipation of it, were described as having sustained psychological consequences, including anxiety and hypervigilance.

Neighbourhood cohesion emerged as a significant factor in shaping perceptions of safety and mental well-being. Maria contrasted her experiences across two boroughs:

> *I’ve never felt so unsafe in Harrow and the reason why is because in Lambeth … people look out for each other. There’s always that feeling that it’s okay, we’re in it together. But I didn’t feel that over there [in Harrow]. And to the opposite I felt … “we’re not helping you, you’re not from our community, you’re not from our group” … oh I’m so lucky at least where I am we look out for each other, even if we don’t look the same, we don’t talk the same, we don’t pray the same, we look out for each other, you know. – Maria, 45-54, White other*

Maria’s account demonstrates that her sense of safety was shaped by the presence (or absence) of mutual care within the neighbourhood. In Lambeth (a borough in south London), she experienced a shared responsibility among residents, which reduced her feelings of vulnerability. In Harrow (a borough in northwest London), she felt positioned outside of that collective, which heightened her sense of exposure and unease. Maria’s narrative shows that diversity within the community did not weaken her sense of security. People differed in social demographics, yet she described feeling protected where everyday recognition and support were visible. Being included within a network of care eased fear and strengthened mental well-being, whereas exclusion intensified insecurity.

Similarly, Nadine emphasised the role of neighbourly attentiveness in shaping her sense of security:

> *The impact that my community have on me is, number one, safety, I feel safe in my community because I know my neighbours always watching out for me at any given time. So that’s what it has done for me. – Nadine, 35-44, Black Caribbean*

Within this account, safety is situated in the awareness of being seen and cared for by others. Knowing that neighbours are attentive to her presence conveyed reassurance and reduced underlying anxiety. This attentiveness was expressed through everyday awareness and oversight, creating a sense of support within the neighbourhood. This ongoing, everyday vigilance reduced anticipatory fear. Participants linked this sense of relational awareness to improved mental well-being, describing how knowing others were invested in their safety strengthened stability and their ability to cope with potential risks.

However, participants also recognised the fragility of these protections when structural conditions undermine them. Tony reflected on how environmental deficiencies shaped his sister’s anxiety:

> *She said she’s got really poor streetlighting in her area. She hasn’t got intercom system or whatever. That makes her feel terrible, because she’s got two young children … I can empathise with what she’s saying because community support is less proactive and more reactive. And that’s kind of a bad thing. – Tony, 45-54, Black Caribbean*

Tony’s narrative illustrates how institutional neglect amplifies psychological distress. Poor lighting and the absence of secure entry systems were described as ongoing sources of anxiety that heightened everyday feelings of vulnerability. Tony’s observation that community support is “less proactive and more reactive” signals the limits of informal reassurance in preventing harm. While neighbours may respond after incidents occur, they cannot substitute for preventative structural measures. As a result, individuals remain in a state of heightened vigilance.

Taken together, these findings indicate that experiences of safety are deeply embedded in social and structural contexts. Neighbourhood cohesion and relational solidarity can buffer anxiety and enhance emotional well-being, while infrastructural deficiencies and exposure to violence intensify vigilance and psychological burden. Safety, therefore, functioned as both a social and mental health determinant, shaped by community inclusion and structural conditions.

### Gendered experiences of safety and responsibility

Gendered experiences were central to how participants navigated everyday risks of violence. One core finding highlighted how women and girls develop daily strategies within their informal networks to manage safety threats, including fear of violence and abuse, walking alone, stalking, and harassment. These strategies to mitigate threats, whilst effective, were sometimes deemed exhausting. For example, Priya adopted a range of safety strategies such as hypervigilance, avoidance, and drawing on trusted informal support:

> *If I do want to go out and leave the house after dark I will try to make sure that I have someone with me or I will try to avoid dark roads or quiet roads … sometimes I put my keys between my fingers. Or sometimes I will call my mum and just have her on the phone while I am walking if I can’t avoid a dark or quiet area. – Priya, 16-24, Mixed ethnicity*

Participants described this reliance on informal support as both practical and emotionally taxing. Safety was often perceived as a constant mental calculation among women, with the burden on risk management falling on women and their immediate networks instead of formal institutions.

Social context also shaped perceptions of safety. Sofia, for example, expressed heightened vigilance when with female friends, but felt significantly safer when accompanied by their male partner:

> *Me and my friends, who are girls, if we’re out really late we would run home, just because there’s a lot of cars and sometimes they will stop. And there’s a lot of random things that make us feel unsafe. But with boyfriend, I just feel quite safe so I don’t really have any precautions. – Sofia, 16-24, Chinese*

These experiences illustrate how informal male protection was often perceived as offering safety, highlighting gendered dynamics in how safety risk is mitigated. Male participants in the study also provided perspectives on safety and gender norms. Kevin described how male-dominated spaces can feel threatening for women:

> *I came to the party here on Friday, there was zillions of women all dressed up … So men are puffing up their chests, sentient beings are puffing up their chest, they want to kind of, you know? And some females may take offence by it, so if they take offence by it, they’re not feeling safe, so you can say for that example this place could be not safe that night. – Kevin, 45-54, Chinese*

Kevin’s account suggests that male behaviour is not necessarily seen as threatening unless interpreted that way by women. This burden of recognising risk is thus placed on women, while the behaviour itself, such as performative masculinity, is left unchallenged. Kevin also acknowledged informal community efforts to support female victims of violence, often organised by women, but positioned himself as peripheral to these initiatives:

> *I have a few friends that live within my community, they run these organisations where I don’t get involved. I don’t get involved in it but I know they exist, so rape victims, you know, torture victims, female. So I know it exists, I know that it happens. How I support them? I don’t. I just let them get on with [it]. – Kevin, 45-54, Chinese*

Kevin’s account illustrates that while some men may express admiration for the women leading efforts in tackling VAWG, they also maintain a distance, revealing a gendered division in emotional labour and responsibility for community safety.

In addition to in-person networks, digital platforms also operated as key sources of informal support among women. Women used social media to raise awareness and identify threats, as Aisha explains:

> *I think it plays a big role, because I feel there are a lot of circumstances where sometimes women don’t even know that they’re being abused. And I feel social media brings to light things that you wouldn’t necessarily be aware of. It’s a way of education and sharing knowledge, and then it’s also a way of bringing light to resources and ways that you can receive help. – Aisha, 16-24, Black African*

Through these platforms, women obtained access to valuable safety resources and shared knowledge, strengthening informal support networks and empowering individuals to recognise and respond to safety threats in the community and beyond. This was further reinforced by Sofia’s experience, with them sharing how women use TikTok and Facebook to alert each other about potential danger locally:

> *Girls support girls, so on TikTok, I think sometimes girls give advice to other girls in areas where they live or on Facebook, it’s group chats with only girls and they might alert each other about some dangerous people or something that happened in the local area. – Sofia, 16-24, Chinese*

These digital networks offered immediacy and collective care. They also served as compensatory spaces where women validated each other’s experiences in ways often lacking in formal services.

Across this theme, narratives highlight the mental and emotional burdens associated with safety work. While informal networks offered information and solidarity, gendered expectations positioned women as primarily responsible for vigilance. Although these networks could buffer the emotional impact of violence, they simultaneously reinforced traditional gender roles.

### Formal support and barriers

Participants described a range of overlapping barriers that limited their ability to draw on formal health systems to address safety needs. Although such services were technically available, women in particular explained they were effectively inaccessible due to a combination of institutional and cultural barriers. These included the complexity of navigating services, lack of awareness, stigma, as well as fears of being misunderstood. As a result of these barriers, a reliance on informal support networks emerged yet again, however, these too were not always consistently available, leaving some individuals without these networks particularly vulnerable to distress.

A general concern among participants was that formal systems were difficult to approach because they were complex, and how this contributes towards fear and isolation:

> *Probably, just not knowing the processes, they’d just probably be scared. You feel unsafe … you’re not going to want to be around other people and socialise. You might want to isolate more. But, also, you know, your social bonds might help you. Those might be the people that you can find and help you in your situations … it could be a phone call. It could be someone accessing a number for you. Setting up a meeting. – Deborah, 35-44, Black Caribbean*

Deborah’s reflection showcases how navigating formal services often depends on others acting as intermediaries. Where social bonds are weak or absent, participants described how many fall through institutional gaps, resulting in prolonged distress and an increasing reliance on avoidance as a coping strategy. Yasmin expands on another issue when limited community awareness of services acts as a barrier:

> *Well, I suppose women need to feel that there are safe spaces in their community that they can go to. I think in my community we have a centre and there’s that provision for a lot of things for females there so that’s a safe space. So, we do have those safe spaces but then again, it’s about awareness, isn’t it? – Yasmin, 45-54, Black African*

This lack of visibility meant that formal routes to safety and emotional support were, in practice, inaccessible to those most marginalised. Other participants reflected on cultural norms that discourage seeking help externally. Isabella explained how internalised expectations of self-reliance and a desire for privacy can prevent individuals from disclosing harm outside the family:

> *I feel like it’s like a cultural thing* … *A lot of people are taught to be independent and any issue that happens should just stay between your family, like it’s not anyone’s business really. So, I feel like that could stop a lot of people from going out to speak to people and they’re not sure what they can get out of it. – Isabella, 25-34, Black Caribbean*

Priya further highlighted the stigma surrounding family or sexual violence in certain communities:

> *I think they are less likely [to seek help] because … there is more of a stigma at least in Indian culture around family violence and sexual violence … it is taboo to talk about it in the first place. But then to go and seek support for that there is going to be less of an assumption that people are going to understand you. Whether there is language barriers or cultural barriers there might be more of an assumption that you are just not going to be understood so what’s the point? – Priya, 16-24, Mixed ethnicity*

Such accounts made clear that cultural taboos and fears of confidentiality breaches created emotional barriers that prevented women from seeking help, even when experiences of harm were significant. Shame or the threat of community backlash often produced silence, which participants linked to increased distress and resignation.

These challenges were intensified for those with migration-related vulnerabilities. Julie explained how newcomers often struggle because they lack knowledge of expected procedures of interacting with formal institutions:

> *If you’re not aware of it people fall through the cracks, and that’s really sad because you don’t know how to access it. So, I do think that these organisations, there is an additional duty or, just understanding that they have to have for people from different minorities that aren’t used to how the system runs … do we provide an environment that makes everyone feel that it’s safe and that they’re able to access things? No, so I think a lot of work has to be done to make sure that they do feel that. – Julie, 35-44, Black African*

Language barriers, unfamiliar bureaucratic processes, and fear of institutional authority left many migrant women unsure of whether seeking help might expose them to further harm. Participants suggested that such structural forms of exclusion contribute to prolonged uncertainty and a deep sense of being unprotected.

Across accounts, participants repeatedly noted that the perceived cultural insensitivity of formal mental health services further discouraged engagement. Several women assumed in advance that their experiences would not be understood or taken seriously because their cultural contexts differed from those of service providers. Priya captured this sentiment:

> *I think they are always just going to be less likely to access them because there’s no real sense of intersectionality when it comes to helping marginalised people from a greater issue, for example sexual harassment, that’s a problem for everybody. But there’s definitely a lot more taboo and issues when it comes to sexual harassment of someone of an ethnic minority* … *some people may not go to these kinds of groups out of fear of shame. Out of fear of it getting out or something, if there’s a lack of confidentiality. – Priya, 16-24, Mixed ethnicity*

The expectation of being misunderstood thus functioned as a major barrier. Even before any interaction occurred, participants anticipated judgement or minimisation of their experiences. As a result, many avoided formal support, a pattern that in turn compounded their distress.

These barriers also shaped how participants relied on informal networks. While family and friends often played important roles in helping women interpret or navigate formal systems, reliance on such networks led to inconsistent levels of support. Those without reliable social ties, or whose communities upheld strong norms of silence, described being left alone to manage safety concerns and emotional suffering. Some participants also noted that informal networks, such as ties within community groups, could reinforce stigma or discourage disclosure, meaning that the spaces women turned to for refuge could simultaneously limit their options for seeking help. The result was that women who were most isolated due to migration or community expectations were also those most likely to face worsening mental health impacts when formal systems felt inaccessible.

Taken together, narratives within this theme suggest the effectiveness of formal systems cannot be fully understood without considering the informal support networks and cultural dynamics that surround them. Participants detailed how they frequently relied on friends, family, and the wider community for emotional and practical support in navigating (or avoiding) formal systems. However, when trusted networks were absent, or were themselves bound by stigma, participants expressed how women in particular may be forced to endure their struggles in silence rather than risk the consequences of seeking help. Such accounts pointed to the emotional burden that silence can impose, with participants suggesting that in the absence of supportive networks, women might experience heightened distress and shame when left to cope alone.

### Community and peer-led initiatives as a response to institutional distrust

Across interviews, participants described the emergence of community and peer-led initiatives as a direct response to institutional distrust. These initiatives were viewed as necessary alternatives where public safety institutions were perceived as unreliable or emotionally unsafe.

Distrust in institutions was rooted in both lived and collective experiences. Participants described policing systems as sites where protection and harm coexisted, producing ambivalence. Sarah’s account was particularly illustrative of this tension:

> **Interviewer: *What does long-term support around safety look like for you?***
>
> *To be honest, I don’t know. Because, I say as a Black woman I would like to see more police, but what does that mean for young Black boys who are disproportionately targeted? So, I can’t really say the police. You know, once upon a time they used to say the police was your friend, that used to be a whole marketing strategy. And I know that not to be true* … *It’s only community groups that can help. – Sarah, 35-44, Black African*

Sarah’s narrative captures a central finding, in that participants often desired safety but distrusted formal systems. The question “what does that mean for young Black boys?” reflects how safety is evaluated collectively. Institutional protection for one group member was perceived as potential harm for another. This racialised dilemma produced uncertainty around state-based solutions and repositioned community groups as the only viable source of protection.

This distrust also shaped immediate decision-making. Ifeoma described the emotional tension involved in deciding whom to contact in moments of risk:

> *It probably wouldn’t be the first phone call I make. I would call my mum first before I call the police. I feel there’s a general aura of distrust, and a lot of the time they abuse their power. And, unless it had absolute kind of concrete or conclusive evidence, I wouldn’t feel happy interacting with them. If I went to my mum, she would give me more a response that has more empathy in it, and she’d probably know what to do anyway. I feel safer with my mum than I would with the police. – Ifeoma, 16-24, Black African*

Here, the threshold for institutional engagement is high. The need for “conclusive evidence” suggests that participants anticipated scrutiny or dismissal. Psychological safety, defined by empathy and trust, was described as more readily available within informal networks. Turning to her mother reflected confidence in a familial response grounded in care. Informal relationships were therefore described as psychologically protective spaces, reducing anticipatory anxiety associated with institutional contact.

Participants also described more organised forms of community-led safety initiatives that operated outside immediate family networks. Jason referred to vigilante groups in Nigeria as an example of collective responsibility for protection:

> *In Nigeria we have vigilante groups where people would volunteer to go out at night to patrol the local area to deter crime. I think that’s a good option that we could adopt in local communities here. There were just ordinary members of the community volunteering to protect the community from crime at night. – Jason, 45-54, Black African*

Jason’s emphasis on ‘ordinary members of the community’ reflects protection situated within participatory dynamics and embedded in local accountability. Community patrols are presented as pragmatic responses to perceived institutional insufficiency. Similarly, Aisha described a Black-led grassroots organisation responding directly to racial abuse in her community:

> *There’s this organisation … this black power organisation that exists. And they’re a community initiative. They have a social media page, but there’ll be incidents in the community. There was an incident where a boy was racially abused in our community, and they went, they’ll sort it out. So, I think there’s a rise of these community-started initiatives, and people turning to them instead of the police. I think what they’re doing is really, really important, and it’s really needed. And I like that they’re kind of changing the narrative. – Aisha, 16-24, Black African*

Aisha’s account highlights two interrelated dynamics. First, community initiatives are described as immediate responders to racial harm. Second, these initiatives are framed as “changing the narrative.” This suggests that community-led responses reshape how racialised violence is publicly understood. By responding collectively, these groups counter feelings of invisibility, enhancing communal confidence and solidarity.

Across accounts, participants described community and peer-led initiatives as culturally attuned and emotionally responsive in ways formal systems were not. Grounded in shared experience, these initiatives reduced fear of misinterpretation. At the same time, participants emphasised that they developed in response to unmet needs within formal systems. The reliance on informal enforcement thus reflected perceived institutional failure. While described as protective, these initiatives operate outside the formal accountability structures that govern statutory systems. Jason’s example, for instance, did not specify how authority is regulated within such vigilante groups. The power to define and enforce “safety” remains internally situated, raising implicit questions about accountability.

Overall, the data within this theme indicates that community and peer-led initiatives function as alternative safety infrastructures within racially and ethnically minoritised communities. They enhance perceptions of safety through relational trust, shared responsibility within the community, culturally informed responses to harm, and the immediacy of locally embedded support. At the same time, their emergence reflects enduring institutional distrust and the emotional strain of navigating systems perceived as racially inequitable. These initiatives thus signal both collective resilience and dissatisfaction with formal systems.

## Discussion

The study explored the role of informal support networks in shaping responses to community safety and mental well-being among racially and ethnically minoritised groups. Thematic analysis of findings identified key themes: (1) experiences of community safety and their mental health impacts, (2) gendered experiences of safety and responsibility, (3) formal support and its barriers, and (4) community and peer-led initiatives as a response to institutional distrust.

### Community safety and mental health impacts

Consistent with wider research linking neighbourhood cohesion to better mental health outcomes (Fone et al., 2014), participants described feeling safer and less anxious when embedded in supportive local networks. However, the findings also critique the idea that cohesion is simply protective. Participants linked safety directly to material conditions such as lighting and visible community presence. Where infrastructure was weak, anxiety increased, even when neighbours were supportive. This reinforces public mental health research showing that exposure to unsafe environments contributes to poorer well-being (Guite et al., 2006). In other words, informal networks may provide reassurance, but they cannot fully offset structural neglect.

The findings therefore suggest that safety operates at both relational and structural levels. While cohesion can mitigate distress, it does not eliminate the mental health impact of environmental insecurity. This reinforces arguments that community resilience should not substitute for sustained urban investment (McCay et al., 2019).

### Gendered experiences of safety and responsibility

The second theme strongly aligns with literature on violence against women and girls (VAWG), which documents the everyday strategies women adopt to manage risk. The predominance of women’s accounts in this study brought into focus the gendered organisation of safety work. Women in this study described constant vigilance: avoiding dark spaces, walking alone, sharing locations, calling family members, and using digital networks for support. These strategies mirror existing findings that women’s safety work is normalised with significant emotional and practical burdens (García-Moreno et al., 2025).

What is particularly clear here is how this labour is organised through informal networks. Mothers, friends, community members, and online peer groups functioned as immediate safety infrastructures. Social media platforms were used to circulate warnings and increase awareness of abusive behaviour, echoing wider research on digital feminist solidarity (Vachhani, 2024).

At the same time, the findings highlight a persistent gender imbalance. While some male participants acknowledged women’s safety concerns, they did not consistently engage in preventative or support work. This reflects broader evidence that emotional and practical safety labour is disproportionately carried by women, even as men constitute the majority of perpetrators of violence (Jewkes et al., 2015). Informal networks therefore protect women, but they also reproduce gendered expectations that women manage their own risk.

It is important, however, to situate these findings within the gendered composition of the sample. As the majority of participants were women, the analysis foregrounds women’s experiences of community safety. This should not obscure the distinct and well-documented exposure of young men, particularly Black men, to serious community violence and police brutality (Burrell et al., 2021). These experiences may generate different forms of psychological distress, such as hypervigilance and anxiety, as well as institutional distrust. The findings here therefore show one dimension of gendered safety work, while wider literature reminds us that structural racism and violence operate across gender in different but interconnected ways.

### Formal support and its barriers

Participants’ hesitation to engage with formal services reflects well-documented patterns of institutional distrust among racially and ethnically minoritised communities. Consistent with existing research on racialised policing and mental health in the UK, participants described high thresholds for contacting police, often citing mistrust (Black Thrive Global & Psi, 2023). Decisions in this study were shaped by individual experiences and collective concerns about racial profiling.

In mental health research, barriers to accessing mental health services among racially and ethnically minoritised groups are often framed in terms of stigma or lack of awareness (Misra et al., 2021). While these were present in this work, we show that anticipated discrimination and cultural misunderstanding were also significant. Participants feared not being believed or taken seriously, particularly in cases of sexual or domestic violence. This aligns with critiques that formal services frequently fail to account for intersecting racialised and gendered experiences (Hulley et al., 2023).

Where formal services felt inaccessible, informal networks acted as intermediaries, helping individuals locate resources or decide whether to seek help at all. However, reliance on informal mediation meant access depended on personal connections. Those without strong social ties were at greater risk of isolation, which can have detrimental impacts on mental health outcomes (Evans & Feder, 2016).

### Community and peer-led initiatives as responses to distrust

Participants described grassroots organisations, informal patrols, and peer-led interventions as crucial for supporting community safety and mental well-being. These initiatives were perceived as immediate, culturally responsive, and grounded in shared experience, aligning with literature on collective efficacy and mutual aid within marginalised communities (Spade, 2020).

While community-led responses enhanced feelings of safety, they also highlighted the enduring influence of structural racism and violence on protection. In contexts where formal institutions were perceived as harmful, informal systems often assumed primary responsibility for safety. Although highly valued, these initiatives relied heavily on unpaid labour and operated within the same structural conditions that generate insecurity.

### Implications

Improving mental well-being in racially and ethnically minoritised communities requires addressing both environmental insecurity and institutional distrust.

First, neighbourhood infrastructure should be treated as a mental health issue. Targeted investment in public infrastructure and community safety audits could reduce everyday anxiety and hypervigilance described by participants.

Second, responses to VAWG should build on existing informal networks among racially and ethnically minoritised groups. Embedding trauma-informed advocates or drop-in mental health support within trusted community spaces (e.g., community centres, faith settings) may reduce barriers linked to mistrust.

Third, rebuilding trust in statutory services requires visible accountability. In policing, this could include strengthened community oversight mechanisms and transparent review of stop-and-search practices, particularly given concerns about over-policing of young Black men, as described in this study.

Finally, mental health services must respond to the barriers participants described, particularly anticipated cultural misunderstanding, concerns about confidentiality, trust, complex processes, and lack of awareness of services. Simplifying access and strengthening culturally informed trauma support may reduce the need for informal mediation.

### Limitations

A key strength of this study lies in platforming the lived experiences of racially and ethnically minoritised communities and providing in-depth insight into how informal and formal safety networks are negotiated in contexts of structural inequality. However, given its urban focus and predominantly female sample, the findings may not fully represent experiences in other geographical settings as well as adequately capture male perspectives.

## Conclusion

This study highlights the central role of informal support networks in shaping community safety and mental well-being among racially and ethnically minoritised groups in South East London. While these networks provide vital practical and emotional support, they operate within conditions of structural neglect and cannot replace efficient public safety and mental health infrastructures. Viewing reliance on informal systems as a deficit overlooks the fact that it reflects community resourcefulness and resilience. A holistic approach, one that values community-led strengths while investing in accessible formal services, can enable racially and ethnically minoritised groups to shape safety on their own terms while ensuring that robust mental health support is available when needed.

## Data Availability

All data produced in the present study are available upon reasonable request to the authors

## Acknowledgements

We express gratitude to all participants who generously shared their experiences. We also would like to thank our peer researchers for their dedication and insights throughout the work and to our partner organisations for their invaluable support.

